# Wave Reflection: More Than A Round Trip

**DOI:** 10.1101/2020.03.30.20048223

**Authors:** Rashid Afkhami, Sarah Johnson

**Affiliations:** School of Electrical Engineering & Computing, The University of Newcastle

## Abstract

Reflected pressure waves are key to the understanding of vascular ageing, a prominent factor in major cardiovascular events. Several different metrics have been proposed to index the effect of wave reflection on the pressure waveform and thereby serve as an indicator of vascular ageing. The extent to which these indices are influenced by factors other than vascular health remains a matter of concern. In this paper, we use transmission-line theory to derive a mathematical model for the reflection time (T_refl_), and the augmentation index (AI), assuming a general extended model of the arterial system. Then, we test the proposed model against values reported in the literature. Finally, we discuss insights from the model to common observations in the literature such as age-related “shift” in the reflection site, the variation of AI with heart rate, and the flattening of T_refl_ in older participants.

## 1 Introduction

Vascular ageing is a prominent factor in major cardiovascular events including stroke, heart failure and coronary artery disease [1]. Vascular health is studied through pulsatile arterial hemodynamics and several key indicators of vascular ageing have been identified in pulsatile pressure readings such as the pulse wave velocity (PWV), reflection time (T_refl_), and augmentation index (AI) [1, 2].

In particular, reflected waves are frequently studied to infer cardiovascular properties [2]. Reflected waves occur when forward travelling pressure waves hit an effective reflection site (which is a superimposition of several sites in practice) and are reflected back towards their source (in this case the heart). In elastic arteries appropriately timed reflected waves help maintain pressure during diastole, however, they can have an ill-effect as age progresses and vessels stiffen, leading to rise in PWV. With increased PWV, which more than doubles in the aorta between the ages of 17 and 70 [3], reflected waves advance into the systole and add to the systolic pressure [4]. This increases peak systolic, end diastolic pressure and mean arterial pressure [5] and contributes to stress on the vessels [2].

The concept of reflection time, pulse transit time or pulse return time was defined on the central pressure waveform as the timing of the inflection point formed when a forward travelling pressure waveform from the left ventricle combines with a reflected wave [6]. Here, we will refer to this timing as the reflection time, T_refl_. T_refl_ is commonly formulated as

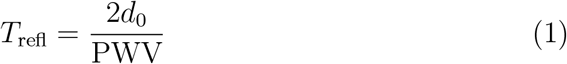

where *d*_0_ is the distance from the measurement site to the reflection site, multiplied by two to account for a round trip. The equation simply calculates the travel time by dividing distance to travel speed. This equation is commonly used to assess vascular compliance or to estimate *d*_0_ [7, 8, 5, 9, 6, 10, 11, 12, 13, 14, 15]. It should be noted that (1) holds only when a resistive load is assumed, i.e., a real load (in a mathematical sense) impedance [6, 15, 16]. However, a pure resistive load can not sufficiently model distal arteries [16]. In elderly populations (>65 years) T_refl_ reaches a plateau state whereas PWV still increases and the first conclusion from (1) is increased *d*_0_, the apparent distal shift of the reflection site [5, 9] which contradicts with the accepted opinion [17]. The debate of changes in *d*_0_ has been the topic of several publications [6, 16, 5, 9]. In this paper will examine the controversy of moving reflection site and other observations for the reported T_refl_ and AI values by understanding these indices from a mathematical viewpoint using a model of the vascular tree.

In this paper, we model the vascular system as a three-element Windkessel (WK3) [18, 19, 20] at the load site and use the transmission line (TL) theory [19, 21] to formulate T_refl_ and AI in terms of the model parameters. Then, we use the reported values in the literature for each parameter and compare measured T_refl_ and AI values to the model outputs. After validating proposed models for T_refl_ and AI, we use them to gain insights into commonly observed measurements.

## 2 Method

### 2.1 Mathematical Models of T_refl_ and AI

The heart is connected to a TL with characteristic impedance of *Z*_0_ and terminated by a WK3 with its third element, *Z*_0_, matching the characteristic impedance of the line, see Fig. 1. *R* and *C* are the resistance and the compliance, respectively, resembling the properties of the vascular system beyond the reflection site and *d*_0_ is the distance between the measurement site and the reflection site.

**Figure 1.**
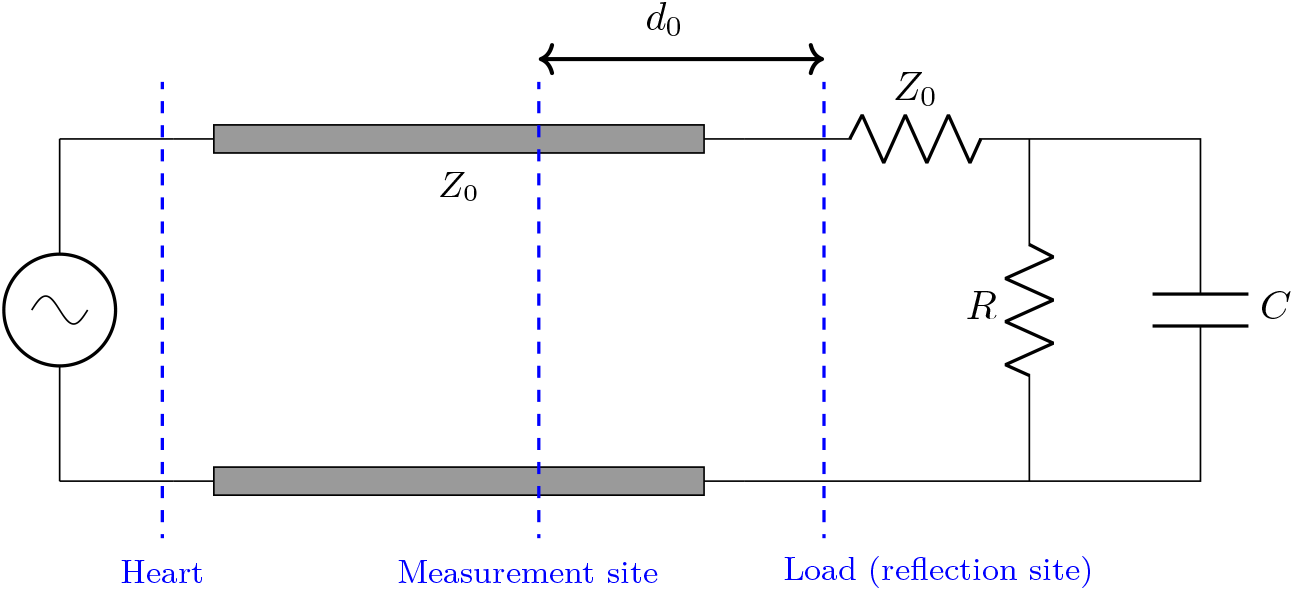
Transmission line model

**T**_**refl**_: Using TL theory we show (see supplementary materials 1) that

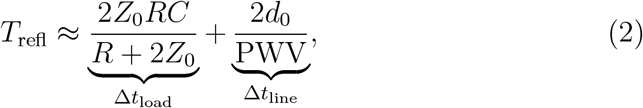

which breaks the travel time of the reflected wave into two elements. Δ*t*_line_ is the delay caused by the line itself which is influenced by the speed on the line and the length of the line. Δ*t*_load_ is the delay at the load, in particular forced by the capacitive properties of the load. Thus, the wave reflection is more than a simple “round trip”, there is a delay in between.

**AI:** The augmentation index, or AI, is commonly defined in distal arteries as:

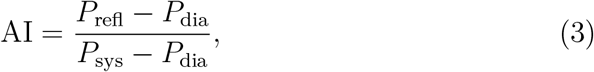

in which *P*_refl_, *P*_dia_ and *P*_sys_ are peak reflection, end-diastolic and peak systolic blood pressures, respectively [22]. For proximal arteries where reflected and the incident waves often overlap, another definition is used.

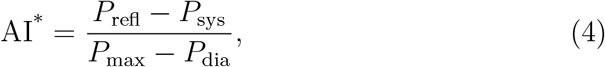

where *P*_max_ = max {*P*_refl_, *P*_sys_} [7, 23, 24, 13, 25]. Using a similar theoretical approach (see supplementary materials 1) we show that

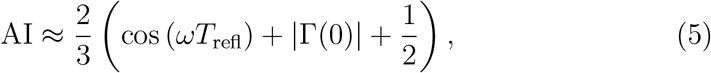

where *ω* is the angular velocity and Γ_0_ is the reflection coefficient at the load site, defined as the ratio of the reflected pressure wave amplitude to the incident pressure wave amplitude. AI^*^ can be calculated using the conversion equation of (6).

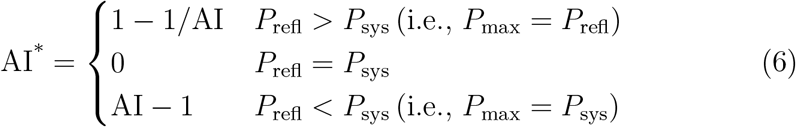

### 2.2 Model Validation

In this section we will compare measured values reported in the literature for T_refl_, AI and AI^*^ with values derived from proposed models. It is important to note that we do not “fit” proposed model to the data. All model parameters are set based on existing known values for vessel properties from the literature. Four different cases are studied which have measurements from the carotid/radial arteries (see supplementary materials 2 for detailed explanation of the vessel parameters used).

- **Case I**: In a large-scaled study by Segers et al. [26], 2026 healthy middle-aged subjects are divided into four half-decade age ranges with mean group ages of 37.5, 43, 48 and 53.5 years. The pressure waveform is measured from the left common carotid artery using applanation tonometry (flow is measured from the aorta using ultrasound). T_refl_ and AI^*^ values are reported for each age group. The vessel parameters for this case are reported in Table 1. We compare the values for T_refl_ and AI^*^ reported by Segers et al. [26] with the values calculated using (2) and (6). We also calculate T_refl_ using the existing model as per (1).
- **Case II**: For this case we use the data of 266 healthy participants (age range of 18-78 years and mean *±* standard deviation (SD) of 37.9 *±* 18.9 years) reported by Zhang et al. [27]. Radial arterial pressure is measured using applanation tonometry, the time interval between systolic and reflected peaks is reported as T_refl_ and we use these values to derive T_refl_ with both (1) and (2). Vessel parameters for this case are listed in Table 2.
- **Case III**: A radial augmentation index is reported by Kohara et al. [22] for 632 healthy subjects, where AI is defined as in (3). We derive model-based AI for the radial artery using (5) and parameters listed in Table 2 and compare with the reported values by Kohara et al. [22].
- **Case IV**: Changes of aortic T_refl_ with PWV in 73 outpatients (age range 17-95 years, mean age 51.8 years) is reported by London et al. [28], where pressure waveforms are recorded with non-invasive methods from the carotid artery and are assumed to be similar to the pressure values in the ascending aorta and central arteries. Reported values are gender-independent and are listed in Table 2. We use these values to derive T_refl_ with both (1) and (2).

**Table 1:**
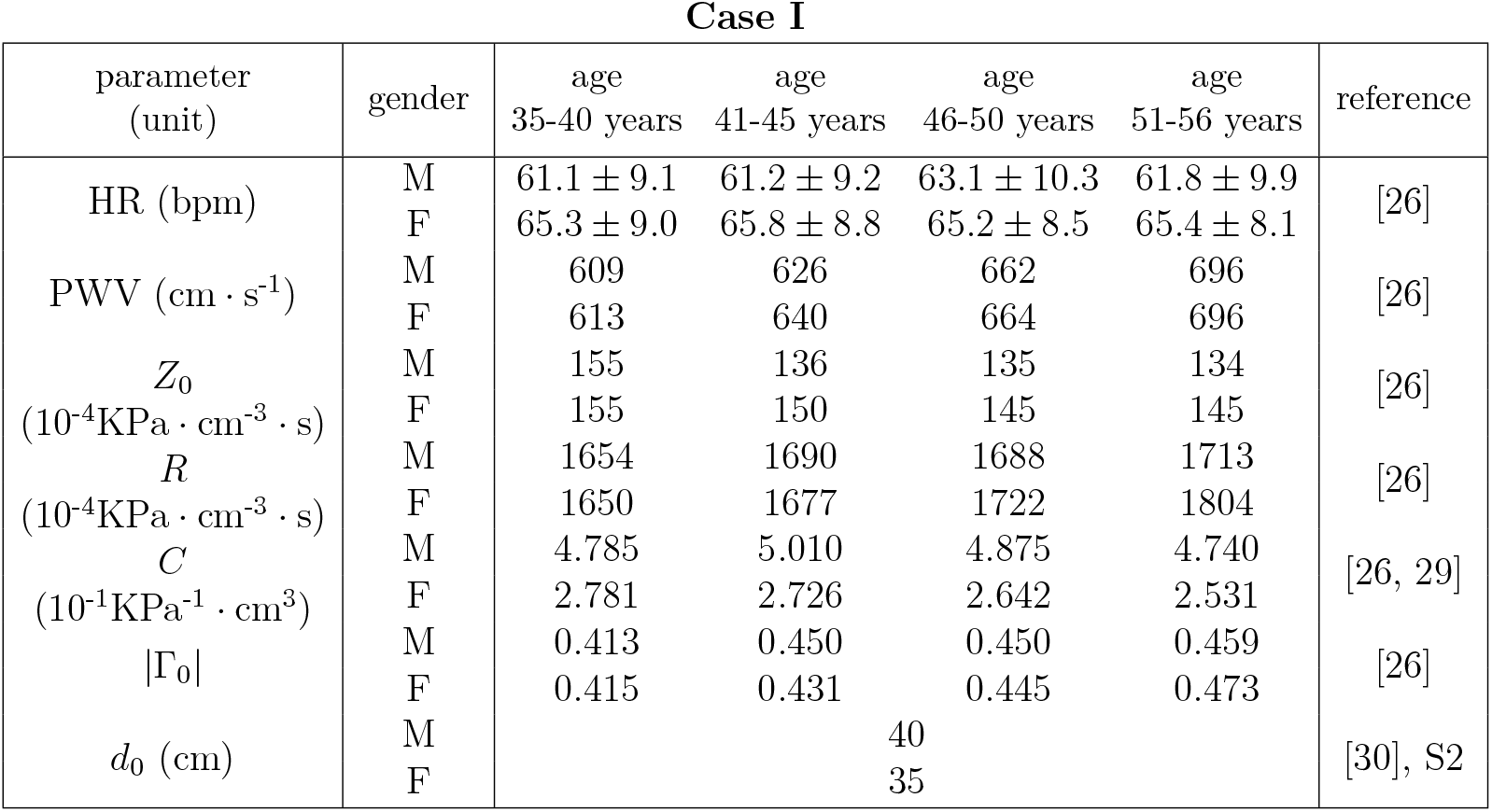
Model parameters for Case I. M: male, F: female, HR: heart rate, S2: supplementary materials 2. HR values are in the form of mean *±* SD

**Table 2:** Model parameters for Cases II, III and IV. M: male, F: female, CA: carotid artery, RA: radial artery, HR: heart rate, S2: supplementary materials 2. HR values are in the form of mean *±* SD.

| Cases II, III and IV |  |  |
| --- | --- | --- |
| parameter<br>(unit) | gender \| artery \| value | reference |
| PWV ($\text{cm} \cdot \text{s}^{-1}$) | $10 \times \text{Age} + 300$ | [31] |
| $R$<br>($10^{-4} \text{KPa} \cdot \text{cm}^{-3} \cdot \text{s}$) | age < 50: $8.1 \times \text{Age} + 926.9 + g_r$<br>age > 50: $333.4 \times \exp(0.0277 \times \text{Age}) + g_r$<br>M \| $g_r = -32$, F \| $g_r = 105$, mixed \| $g_r = 0$ | [29, 32], S2 |
| $C$<br>($10^{-1} \text{KPa}^{-1} \cdot \text{cm}^3$) | $12.39 \times \exp(-0.0277 \times \text{Age}) + g_c$<br>M \| $g_c = 0.77$, F \| $g_c = -0.70$, mixed \| $g_c = 0$ | [29], S2 |
| $d_0$ (cm) | M \| CA \| 40, RA \| 20<br>F \| CA \| 35, RA \| 16 | [33] |
| $Z_0$<br>($\text{KPa} \cdot \text{cm}^{-3} \cdot \text{s}$) | CA \| 0.136, RA \| 1.185 | [34] |
| $\Gamma_0$ | $R / (R + 2Z_0 + 2j\omega Z_0 RC)$ | S2 |
| HR (bpm) | M \| $70.6 \pm 11.0$<br>F \| $71.5 \pm 9.5$ | [22] |

## 3 Results

For cases I-III the results are shown in Fig. 2 and Fig. 3 and for case IV the results are in Fig. 4. Note that Segers et al. [26] reports AI^*^ in the form of mean and standard errors of mean which are converted into mean *±* SD for case I. Error bars for estimated AI in case III are due only to heart rate variability. The results show high similarity between modelled T_refl_ and AI and the corresponding measured values and a high level of improvement for the estimation of the reflected wave compared to the existing model, especially in the radial artery.

**Figure 2.**
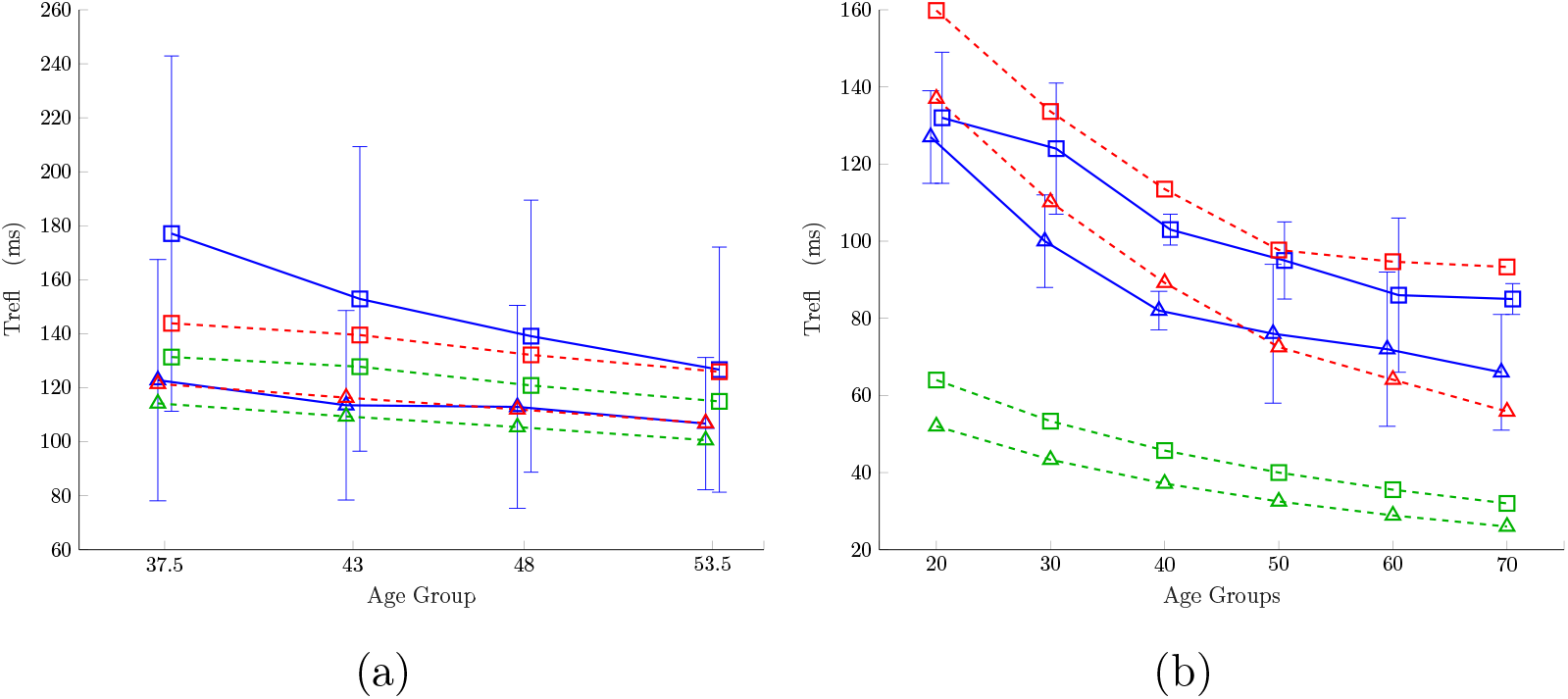
T_refl_ in carotid (a) case I and radial (b) case II for men (square) and women (triangle). Measured (blue, solid) data is from the literature [26, 27] and estimated values are from the new proposed model (red, dashed) and the existing model (green, dashed). Values are in the form of mean *±* SD.

**Figure 3.**
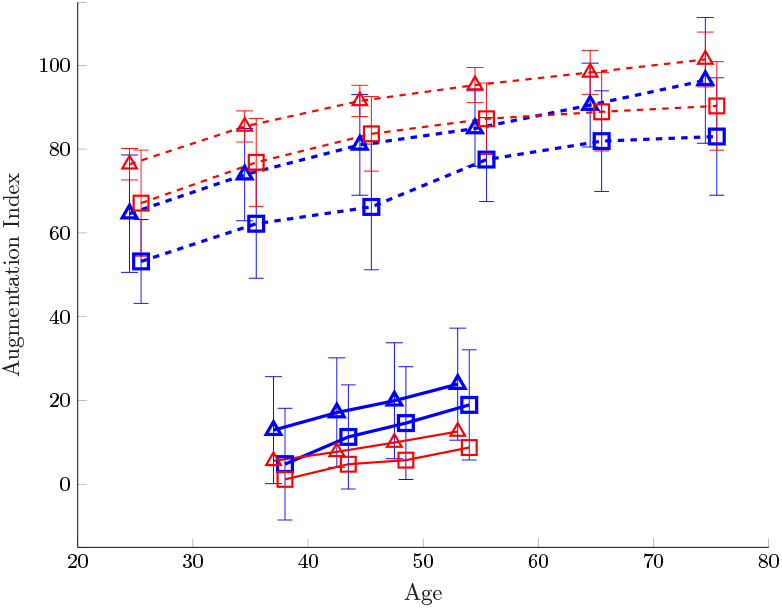
Measured (blue) and estimated (red) augmentation index for men (square) and women (triangle) in carotid (solid line, case I) [26] and radial (dashed line, case III) [22] arteries. Values are in the form of mean *±* SD.

**Figure 4.**
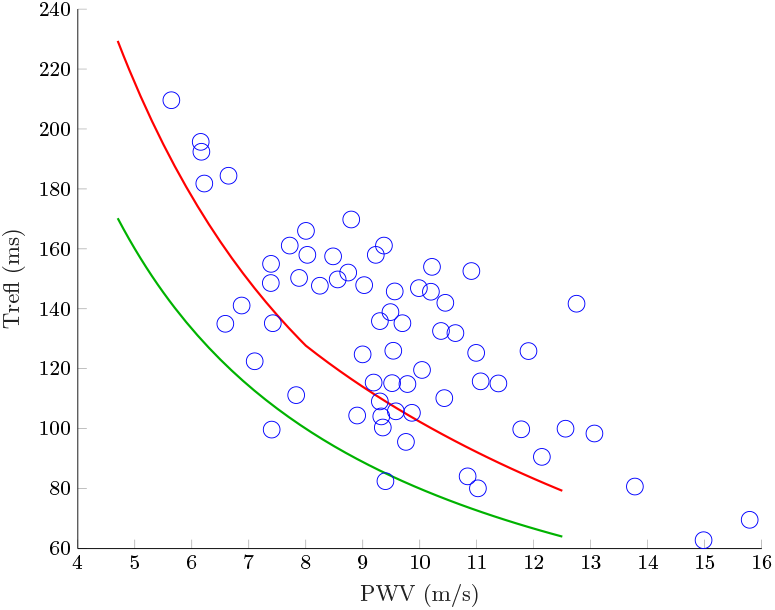
Estimated carotid T_refl_ against PWV using the proposed model (red) and the existing model (green) compared with measured values from London et al. [28] (case IV) in blue.

For case I, linear interpolation of Fig. 3 suggests an age of 34 (men) and 24.9 (women) for AI^*^=0 which aligns with the carotid AI^*^ zero crossing at the ages of 31.7 years for total of 38 male and 18 female participants by Hirata et al. [35]

## 4 Discussion

In this paper we have formulated a mathematical model for T_refl_ and AI including dependencies on both line and load properties. Through several case studies we have compared proposed models for T_refl_ and AI against published measured data, and have shown that they closely match the observed data. We can thus apply these models to give insights into several observed phenomena which we discuss in this section.

Several studies have reported a strong correlation between T_refl_ and age [5, 10, 27]. Based on proposed model we can see that the load site delay of the reflection time, Δ*t*_load_, is primarily influenced by the compliance of the load, as the resistance properties, *R* and *Z*_0_, are found in both numerator and denominator of the model in (2) and so cancel out to some extent. In addition, PWV itself is a function of the line compliance (the compliance of the arteries between the measurement site and the reflection site, commonly used as an index of vascular compliance [36]) and combined with the influence of load compliance (the WK3 compliance at the load) makes T_refl_ a strong index of overall vascular compliance with only weak effects from vascular resistance. It is well known that compliance decreases with age; therefore, the correlation of the reflection time with age is due to strong dependence on changes in compliance with age.

It is reported in the literature that T_refl_ does not decrease linearly with age in older populations and in fact it almost flattens after the age of 65 years [5, 6, 16]. The flattening effect is reflected in proposed model, see Fig. 2, through three main components. First, based on the data reported by McVeigh et al. [29], load compliance does not decrease as sharply with age in older populations (see Table 2). Second, vascular resistance, *R*, increases exponentially after the age of 50 years [32] (see *R* in Table 2), which contributes to an increase in Δ*t*_load_ and thus T_refl_. The third component is the influence of characteristic impedance, *Z*_0_, making the effect highly dependent on the sight of measurement. Based on our estimates from the data by Willemet and Alastruey [34] the value of *Z*_0_ increases almost ten-fold moving from the carotid to radial artery. The small *Z*_0_ values in proximal arteries heighten the flattening effect in T_refl_. Thus, based on proposed model, the flattening of the reflection time is due to reduced changes in compliance and increased changes in vascular resistance with age with the effect more noticeable in proximal arteries due to the decreased impedance in large vessels.

It has been reported that the effective reflection site moves distally after the age of 65 years [5, 9], although this view is challenged in several studies [6, 16]. Our model is able to explain this phenomenon by shedding light on the delay that the reflected wave sees at the load site, Δ*t*_load_. Note that the Δ*t*_load_ portion of T_refl_ is usually ignored in the analysis [7, 5, 9, 13, 14, 8, 6, 16], despite the hints given by Westerhof and Westerhof [37] and Westerhof et al. [16]. Based on proposed model Δ*t*_load_ can account for up to 25% (case IV) and 62% (case II) of T_refl_ in carotid and radial arteries, respectively. Δ*t*_load_ gets larger relative to Δ*t*_line_ when moving distal from the heart for two main reasons; the decrease of *d*_0_ and the increase of *Z*_0_.

It is reported in the literature that reflection time does not show a notable change with heart rate, only 10 ms drop in radial T_refl_ is reported for heart rate ranging from 60 to 80 beats per minute for the data in case II [27]. Our model confirms the independence of heart rate and T_refl_ as heart rate does not affect the calculated T_refl_ in (2).

The augmentation index has been shown to have negative correlations with T_refl_ [38] and heart rate [39] and positive correlation with age [22, 35, 9]. Our model also shows these effects and describes AI, see (5), as a function of T_refl_, heart rate and reflection coefficient. Even in extreme cases, max(*T*_refl_) *<* 0.25 s and max(*f*_HR_) *<* 0.5 Hz (*f*_HR_ being heart rate frequency) and therefore, 0 *< ωT*_refl_ *< π/*2 suggesting that the cosine function is monotonically decreasing and AI will always increase with decreased T_refl_ or decreased heart rate, explaining the negative correlation. Thus proposed model suggests that the positive correlation of AI with age is due to decreased T_refl_ with ageing and the negative correlation between AI and heart rate is due to the presence of the cosine function.

A flattening effect with age is repeatedly reported for the augmentation index [12, 9, 24] after the age of 55 years [24] and can even decline thereafter [9]. Although no decline in AI/AI^*^ was observed in the proposed model, the flattening is noticeable in model outputs with only 1.3% and 2.8% increase in radial AI (case III) between the ages of 65 and 75 years for men and women, respectively. Model suggests that the AI/AI^*^ flattening is due to the flattening of the reflection time, since the other influencing element of the augmentation index is the reflection coefficient which increases, only slightly, with age [26]. Thus, proposed model suggests the observed flattening in AI/AI^*^ is due to age related increase in vascular resistance and more importantly, a slowing of the decrease in compliance with age.

The proposed model for T_refl_ and AI can facilitate studies of vascular ageing by providing means to understand how changes is vascular properties such as *R* and *C* are reflected in these indices [40].

The proposed model contains a series of theoretical assumptions (listed in supplementary materials 1) that form simplified relationships between desired vascular ageing indices and comprehensible model elements, and is subject to practical limitations (listed in supplementary materials 2) which hinder the model validation.

## 5 Conclusion

The proposed models for T_refl_ and AI explain the dependency of both indices on vascular ageing indicators: compliance, PWV and systemic vascular resistance. The model was able to explain the flattening of T_refl_ repeatedly reported in the literature, due to and exponential increase in vascular resistance as well as the moving of the reflection site due to the delay caused at the load by the compliance at the reflection point. Overall, our results suggest that T_refl_ is strongly influenced by vascular compliance and represents a useful index of vascular compliance in populations younger than 65. AI is itself inversely dependent on T_refl_ so shows a similar flattening with age, however it is also strongly affected by heart rate, which will influence AI values independently of vascular compliance.

## Data Availability

NA

