## Supplementary Materials 1 for "Wave Reflection: More Than A Round Trip"

#### 1 Model Derivation

Our aim in this section is to derive a mathematical formula for $T_{\text{refl}}$ and AI using transmission line (TL) theory. We will start with a brief review of the TL theory. The approach is based on a uniform TL model of the vascular system used in the literature [1, 12], see Fig. 1. The heart is located at $x = -d$ with blood pressure and flow of $P_{\text{H}}$ and $Q_{\text{H}}$, respectively. $Z_0$ is the characteristic impedance of the transmission line and the reflection site has the blood pressure and flow of $P_{\text{L}}$ and $Q_{\text{L}}$, respectively terminated by a three-element Windkessel with its third element, $Z_0$, matching the characteristic impedance of the line. $R$ and $C$ are the resistance and the compliance, respectively, resembling the properties of the vascular system beyond the reflection site. It should be mentioned that a tapered model [10] can be used instead of a uniform model, however, given a fixed measurement distance the two models can be shown to be equivalent given an appropriate optimization of the parameters [6]. Note that the TL model accounts only for the pulsatile components of the pressure and flow.

The pressure as a function of time and distance can be decomposed into forward and backward travelling elements, i.e.,

$$P(x) = P_{\text{f}}(x) + P_{\text{b}}(x). \tag{1}$$

In which $P_{\text{f}}(x)$, $P_{\text{b}}(x)$ are the forward and backward (reflected) travelling waveforms. These waveforms are in the form of

$$P_{\text{f}}(x) = p_{\text{f}} e^{-\gamma x}, \quad P_{\text{b}}(x) = p_{\text{b}} e^{\gamma x}, \tag{2}$$

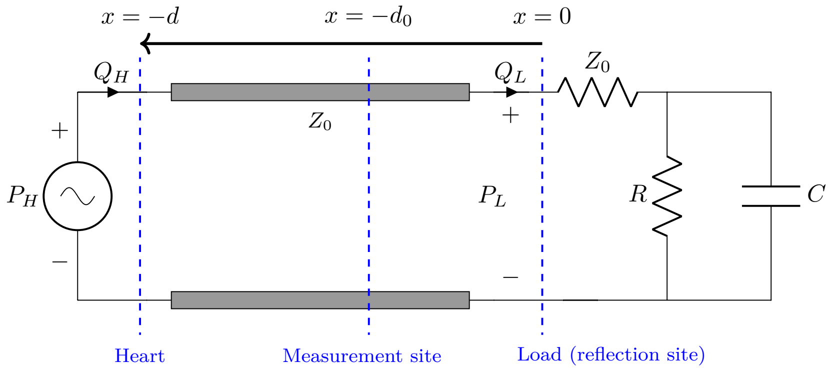


Figure 1: Transmission line model

where $p_f$ and $p_b$ have complex quantities in general and are calculated using the boundary conditions. $\gamma$ and $x$ are the propagation constant and distance from the load, respectively. Note that (1) and (2) are phasor domain solutions to the transmission line equations assuming steady state sinusoidal pressure and flow input waveforms. For a lossless case, the time domain solution will be

$$p(x,t) = |p_{\mathrm{f}}| \cos(\omega t - \beta x + \phi_{\mathrm{f}}) + |p_{\mathrm{b}}| \cos(\omega t + \beta x + \phi_{\mathrm{b}}), \tag{3}$$

where $|p_{\mathrm{f}}| e^{j\phi_{\mathrm{f}}}$ and $|p_{\mathrm{b}}| e^{j\phi_{\mathrm{b}}}$ are the amplitude and phase of $p_{\mathrm{f}}$ and $p_{\mathrm{b}}$ in (2), respectively and $\beta$, which is equal to the imaginary part of $\gamma$, is called the phase constant. The reflection coefficient is defined as the ratio of the reflected pressure wave to the incident pressure wave, i.e.,

$$\Gamma(x) = \frac{P_{\mathrm{b}}(x)}{P_{\mathrm{f}}(x)} = \frac{p_{\mathrm{b}} e^{j\beta x}}{p_{\mathrm{f}} e^{-j\beta x}} = \frac{p_{\mathrm{b}}}{p_{\mathrm{f}}} e^{j2\beta x}. \tag{4}$$

#### 1.1 Reflection Time

In order to formulate the reflection time or $\mathrm{T_{refl}}$ using TL theory, let $\Phi$ be the phase difference between the forward and the reflected waves at $x = -d_0$ along the line, which will be equal to the absolute value of the phase of the reflection coefficient at the same location, $\theta_\Gamma(x = -d_0)$, i.e,

$$\Phi = |\theta_\Gamma(-d_0)| = |\theta_\Gamma(0) - 2\beta d_0|. \tag{5}$$

To calculate $\theta_\Gamma(0)$, first we should quantify the reflection coefficient at the load, $x = 0$. We have

$$\Gamma(0) = \frac{Z_L - Z_0}{Z_L + Z_0}, \tag{6}$$

where $Z_L = P(0)/Q(0)$, see Fig. 1. Also, with a WK3 at the load, see Fig.1, we have

$$Z_L = Z_0 + \frac{R}{1 + j\omega RC}, \tag{7}$$

inserting into (6) gives

$$\Gamma(0) = \frac{R}{R + 2Z_0 + 2j\omega Z_0 RC}, \tag{8}$$

with its phase equal to

$$\theta_\Gamma(0) = -\tan^{-1}\frac{2\omega Z_0 RC}{R + 2Z_0}. \tag{9}$$

Using the first order Taylor series expansion on (9) and inserting into (5) we get

$$\Phi \approx \frac{2\omega Z_0 RC}{R + 2Z_0} + 2\beta d_0. \tag{10}$$

The phase constant, $\beta$, is related to the pulse wave velocity (or propagation velocity) as $\beta = \omega/\mathrm{PWV}$. Also, note that we are interested in measuring the time difference between the forward and the reflected wave, $\mathrm{T}_{\mathrm{refl}}$, which is the phase difference, $\Phi$, divided by the angular velocity, $\omega$. Inserting these into (10) we have

$$T_{\mathrm{refl}} \approx \underbrace{\frac{2Z_0 RC}{R + 2Z_0}}_{\Delta t_{\mathrm{load}}} + \underbrace{\frac{2d_0}{\mathrm{PWV}}}_{\Delta t_{\mathrm{line}}}, \tag{11}$$

which breaks the travel time of the reflected wave into two elements. $\Delta t_{\mathrm{line}}$ is the delay caused by the line itself which is influenced by the speed on the line and the length of the line. $\Delta t_{\mathrm{load}}$ is the delay at the load, in particular forced by the capacitive properties of the load. Thus, the wave reflection is more than a simple "round trip", there is a delay in between.

It should be noted that the reflection site (Fig. 1) in the proposed model is a symbolic reflection location which represents reflections from various reflection and re-reflection sites [2]. Therefore, $d_0$, the distance between the measurement and the reflection sites, does not indicate a specific location on the arterial system with reference to the measurement site and in fact it can have a value which is larger than what one would expect based on vascular structure. The reason is the existence of re-reflection sites which are closer to the heart than the point of the measurement and reflect back the already reflected waves [2]. These waves with much larger $d_0$ also add up to the measured pressure contributing to the reflected pressure waveform.

#### 1.2 Augmentation Index

The augmentation index or AI is defined as

$$\mathrm{AI} = \frac{P_{\mathrm{refl}} - P_{\mathrm{dia}}}{P_{\mathrm{sys}} - P_{\mathrm{dia}}}, \tag{12}$$

in which $P_{\mathrm{refl}}$, $P_{\mathrm{dia}}$ and $P_{\mathrm{sys}}$ are peak reflection, end-diastolic and peak systolic blood pressures, respectively [5]. An alternative definition for AI used in the proximal arteries where reflected and the incident waves often overlap.

$$\mathrm{AI}^{*} = \frac{P_{\mathrm{refl}} - P_{\mathrm{sys}}}{P_{\mathrm{max}} - P_{\mathrm{dia}}}, \tag{13}$$

where $P_{\mathrm{max}} = \max\{P_{\mathrm{refl}}, P_{\mathrm{sys}}\}$ [7, 4, 3, 8, 11]. $\mathrm{AI}^{*}$ can be expressed in terms of AI as

$$\mathrm{AI}^{*} = \begin{cases} 1 - 1/\mathrm{AI} & P_{\mathrm{refl}} > P_{\mathrm{sys}}\,(\text{i.e., } P_{\mathrm{max}} = P_{\mathrm{refl}}) \\ 0 & P_{\mathrm{refl}} = P_{\mathrm{sys}} \\ \mathrm{AI} - 1 & P_{\mathrm{refl}} < P_{\mathrm{sys}}\,(\text{i.e., } P_{\mathrm{max}} = P_{\mathrm{sys}}) \end{cases} \tag{14}$$

It is known that the reflected waves have negligible contribution during the period between end-diastole and peak-systole because of the lossy line properties of the arterial tree. Therefore, with the absence of the reflected wave we can assume

$$P_{\mathrm{sys}} \approx \mathrm{MAP} + |p_{\mathrm{f}}|\,, \tag{15}$$

where MAP is the mean arterial pressure. MAP in practice is closer to the end-diastole than the peak systole, because of the differences in systolic and diastolic durations. For instance the end diastole, MAP and peak systole are reported by Sesso et al. [9] as 77.5, 93.0 and 124.1 mmHg, respectively. Which shows that systolic peak difference to MAP is twice as great as the end-diastolic peak difference from MAP. Here, for the practicality of the formulation we use the same concept, i.e.,

$$P_{\text{dia}} \approx \text{MAP} - \frac{1}{2}\left|p_{\text{f}}\right|. \tag{16}$$

That is, we have matched the theoretical sinusoidal pressure waveform to MAP and systolic pressure values (rather the alternative of matching it to systolic and diastolic pressures) and have used reported values to estimate the diastolic peak. Now, to quantify the reflected peak value, we should be mindful of the phase difference between the forward and backward travelling waves. Based on the approach taken in Section 1.1, when the reflected wave reaches its maximum value, the incident wave drops its amplitude by the factor of $\cos\Phi$ or $\cos(\omega T_{\text{refl}})$. This means

$$P_{\text{refl}} \approx \text{MAP} + \left|p_{\text{f}}\right|\cos(\omega T_{\text{refl}}) + \left|p_{\text{b}}\right|. \tag{17}$$

In which the second term is the amplitude of the forward travelling wave when the reflected peak happens and the third term is the maximum value of the backward travelling wave at the same time. Inserting (15), (16) and (17) into (12) we get

$$\text{AI} \approx \frac{2}{3}\left(\cos(\omega T_{\text{refl}}) + \left|\Gamma(0)\right| + \frac{1}{2}\right), \tag{18}$$

as our formula for AI. $\text{AI}^*$ can be calculated from (18) using (14). Also, note that in this section we are not assuming sinusoidal waveforms except to define $\text{T}_{\text{refl}}$ in (17).

### 2 Model Limitations

The theoretical assumptions used in forming the model are noted here.

The Taylor series expansion in (10) holds only for small values of arctangent argument, $x$, i.e., $|x| < 1$. However, based on published values [5] we have $\max(x) = 0.50$ radian, making this assumption reasonable for our model.

By assuming a lossless transmission line we are not accounting for reductions in pressure as the pressure wave propagates along the vessel, where in reality the reflected waves will lose power travelling towards the heart.

Thus, we may overestimate $|\Gamma(0)|$ in estimation of the augmentation index. Nonetheless, the results are less affected in distal arteries, as $|\Gamma(0)|$ is not a dominant factor in the calculation of AI. First, because $Z_0$ increases and second, the reflection time is shorter and thus $\cos \omega T_{\text{refl}}$ is becoming the dominant factor in AI.
