## Supplementary Materials 2 for "Wave Reflection: More Than A Round Trip"

---

#### 1 Setting Model Parameters

Four different cases are studied in the main text where reported data from the literature is compared with the results derived from the proposed model. Here, we will describe in details how model inputs parameters are chosen for each case. Among the input variables to the model, the parameters $R$ and $C$ are load properties which will be far from the heart and compliant vessels regardless of the measurement site and therefore location-independent values can be selected for these parameters. However, characteristic impedance, $Z_0$ and pulse wave velocity (PWV) are both line properties which can change as the measured location is moved distal from the heart. Here, we will ignore the PWV increase from aorta to radial artery (the farthest artery we will examine) as this is reported to be small [3] and this will not affect the model results. Yet, we will try to match $Z_0$ to its realistic values depending on the artery of interest.

##### 1.1 Case I

The measurements for this case are recorded from the left common carotid artery and the subjects are divided into four half-decade age ranges between 35 and 56 years. The mean and standard deviation (SD) values for heart rate (HR), characteristic impedance (we used the frequency-domain method results), systemic vascular resistance, $R$, and the reflection coefficient, $|\Gamma_0|$, for men and women are already reported for each age group [10], see Table 1 in the main text.

Although total arterial compliance is already reported by Segers et al. [10], we are interested in load compliance values which are much smaller than when the proximal compliant vessels are involved in the calculations. Therefore, we used mean compliance values of the arteries distal to the heart (referred to as oscillatory compliance by McVeigh et al. [8]) measured invasively by McVeigh et al. [8] (4.852 and 2.670 in the units of $10^{-1}\mathrm{KPa}^{-1} \cdot \mathrm{cm}^3$ for men and women, respectively) to downscale the values reported by Segers et al. [10]. That is, the compliance values (calculated with the pulse pressure method) reported for each age group for men were scaled so that the total mean value would be the same as the one reported by McVeigh et al. [8], and the same was done for the values reported for women. The value of $d_0$ will be challenging to estimate, especially in common carotid arteries where the pressure wave may be affected by the reflected waves coming from both downstream arteries and the aorta. The aortic reflected waves have travelled through the thoracic and abdominal aortas approximately 50 cm away from the measurement site. Considering this, we have set $d_0 = 40$ cm for men and $d_0 = 35$ cm for women, which is longer than the length of the common carotid artery and covers an approximate distance from the arch of aorta to the end of internal carotid artery. The values are approximated based on the distance of 38.6 cm calculated from available data [1].

The input parameter values for this case are summarized in Table 1 of the main text and are used to calculated model-based $\mathrm{T}_{\mathrm{refl}}$ and $\mathrm{AI}^*$ to compare with reported values by Segers et al. [10].

#### 1.2 Case II

Case II is based on radial arterial data reported for healthy subjects with age range of 18-78 years [15]. None of the model inputs are reported by Zhang et al. [15] and therefore, for this case we use values reported in other literature as follows. Values of $C$ are measured with invasive methods by McVeigh et al. [8] for 115 healthy volunteers. Although a linear relationship between $C$ and age is derived by McVeigh et al. [8], non-linear changes are noticeable in the scatter plot and thus we digitized the data to fit an exponential function as

$$C = 12.39 \times \exp\left(-0.0277 \times \mathrm{Age}\right) + g_{\mathrm{c}}. \tag{1}$$

Where $g_{\mathrm{c}}$ is a gender correction parameter, set to +0.77 and -0.70 respectively for men and women to satisfy $C_{\mathrm{men}}(\mathrm{Age} = 40) = 4.85$ and $C_{\mathrm{women}}(\mathrm{Age} =$

47) = 2.67 reported by McVeigh et al. [8] for mean age in each group. To model $R$ in healthy volunteers aged less than 50 years, we have used the linear increase reported by McVeigh et al. [8], first line of (2). However, for ages more than 50 years we have proposed an exponential relationship, second line of (2). That is

$$R = \begin{cases} 8.1 \times \text{Age} + 926.9 + g_\text{r} & \text{Age} \leq 50 \\ 333.4 \times \exp(0.0277 \times \text{Age}) + g_\text{r} & \text{Age} > 50 \end{cases}. \tag{2}$$

This is based on a comprehensive blood pressure study of 2036 participants, stating that the estimation of the vascular resistance using mean blood pressure underestimates the actual resistance value at the ages above 50 to 60 years [4]. The exponential factor is set to the similar rate as of the observed exponential rate in $C$, i.e., 0.0277, and the amplitude of 333.4 is to avoid discontinuity at the Age = 50. The gender correction factor, $g_\text{r}$, is set to -32 and +105 respectively for men and women to satisfy $R_\text{men}(\text{Age} = 40) = 1219$ and $R_\text{women}(\text{Age} = 47) = 1413$ [8].

Pulse wave velocity has the form of

$$\text{PWV} = 10 \times \text{Age} + 300 \quad \left(\text{cm} \cdot \text{s}^{-1}\right) \tag{3}$$

as reported by Gozna et al. [5] measured from the ascending aorta and matches the results by Avolio et al. [2]. Finally, we estimated the characteristic impedance as $Z_0 = 1.185$ KPa $\cdot$ cm$^{-3}$ $\cdot$ s with a WK3 fit to the synthetic data provided by Willemet and Alastruey [14]. Based on the results reported by Segers et al. [10], $Z_0$ either does not change with age or the changes are negligible thus we used a constant $Z_0 = 1.185$ for all ages. We also set $d_0 = 20$ cm and $d_0 = 16$ cm, respectively for men and women corresponding to the length of radial artery (mean value of 18 cm has been reported for the radial artery [12]).

The final input values for this case are summarized in Table 1 of the main text and are used to compare measured and modelled radial $\text{T}_\text{refl}$ values.

#### 1.3 Case III

This case looks at radial AI changes with age [6]. Age-independent heart rate values are reported as 70.6 ± 11.0 and 71.5 ± 9.5 (mean ± SD beats per minute) for men and women, respectively [6]. For this case, all other inputs were selected as as per Case II. Summary of all the input parameters can be found in Table 1 of the main text.

#### 1.4 Case IV

This case reports the changes of aortic $\mathrm{T_{refl}}$ with PWV in 73 subjects with age range of 17-95 years [7]. Reported values were not separated by gender and so we use (1) and (2) to obtain gender-independent estimates of $C$ and $R$ with $g_\mathrm{c} = 0$ and $g_\mathrm{r} = 0$, respectively. We set $d_0 = 40$ cm and $Z_0 = 0.136$ $\mathrm{KPa{\cdot}cm^{-3}{\cdot}s}$, estimated for the carotid artery using a WK3 fit on the synthetic data of Willemet and Alastruey [14].

### 2 Limitations

The practical limitations for this study are listed here. In practice, the calculation of the peak-to-peak time difference between forward and backward waves is difficult, especially in proximal arteries where forward and backward waves do not leave separate peaks. Therefore, various methods have been developed for calculation of the return of the reflected waves. Reflection time is calculated by Segers et al. [10] (case I) using the $4^{\mathrm{th}}$ derivative method [13], referred to as the "shoulder time", $T_\mathrm{sho}$ by Segers et al. [11] whereas London et al. [7] (case IV) use the time of the occurrence of the inflection point, $T_\mathrm{inf}$, as described by Segers et al. [11]. Wave separation analysis can also be used to calculate the time difference of the forward and backward waves using the zero-crossing point of each waveform, called $T_\mathrm{f\text{-}b}$ by Segers et al. [11]. The theoretical approach in this paper uses the peaks of the forward and backward waves to calculate $\mathrm{T_{refl}}$, which is different to the definition of $T_\mathrm{f\text{-}b}$ as each wave shows different rise time from zero-crossing to the peak. However, the modelled $\mathrm{T_{refl}}$ best matches the definitions of $T_\mathrm{sho}$ and $T_\mathrm{inf}$ by Segers et al. [11].

The study was limited by the reported measurements of model input parameters. Especially, age related increase of systemic vascular resistance was reported only in a small number of papers. Although a linear increase of $R$ with age is reported by Nichols et al. [9] and McVeigh et al. [8], it is suggested that $R$ values are underestimated in older subjects [4]. More investigation is required to accurately model changes in $R$ with age.
